# AI4CoV: Matching COVID-19 Patients to Treatment Options Using Artificial Intelligence

**DOI:** 10.1101/2020.11.29.20240614

**Authors:** Andrew I. Hsu, Amber S. Yeh, Shao-Lang Chen, Jerry J. Yeh, DongQing Lv, Jane Y.-J. Hsu, Pai Jung Huang

## Abstract

We developed AI4CoV, a novel AI system to match thousands of COVID-19 clinical trials to patients based on each patient’s eligibility to clinical trials in order to help physicians select treatment options for patients. AI4CoV leveraged Natural Language Processing (NLP) and Machine Learning to parse through eligibility criteria of trials and patients’ clinical manifestations in their clinical notes, both presented in English text, to accomplish 92.76% AUROC on a cross-validation test with 3,156 patient-trial pairs labeled with ground truth of suitability. Our retrospective multiple-site review shows that according to AI4CoV, severe patients of COVID-19 generally have less treatment options suitable for them than mild and moderate patients and that suitable and unsuitable treatment options are different for each patient. Our results show that the general approach of AI4CoV is useful during the early stage of a pandemic when the best treatments are still unknown.

## Introduction

With the rapid increase of coronavirus disease (COVID-19) cases worldwide, effectively treating infected patients is crucial to slow the spread of the disease. However, not all patients are suitable candidates for the most promising treatments. For example, Remdesivir, one of the current approved therapeutic interventions for COVID-19,^1,2^ may not be suitable for patients with pre-existing liver conditions because in vitro studies with liver cell culture systems showed that human hepatocytes are susceptible to Remdesivir-mediated toxicity.^3^ Therefore, these patients are at a high risk of yielding poor outcomes and require other effective interventions.^4^

As of 15^th^ October 2020, there are 3,611 trials for COVID-19 registered at ClinicalTrials.gov. Many of these trials are interventions that have already passed safety regulations or are repurposing drugs already approved for other conditions, and thus may be used to treat COVID-19 patients. A trial’s eligibility criteria, which are usually designed to prove safety and efficacy of a drug for a target population,^5^ can be useful for treatment option consideration. For example, the eligibility criteria of a clinical trial on Remdesivir excludes patients with “severe liver disease”. Accordingly, healthcare teams can exclude Remdesivir for patients with pre-existing liver disease and consider other options.

However, due to the sheer numbers of trials and patients during the pandemic, as well as inconsistent wording in clinical trial criteria,^6^ it is challenging for healthcare teams to assess each candidate clinical trial for every patient. To assist them, we developed a system called AI4CoV (AI for COVID) that can rapidly read the eligibility criteria in ClinicalTrials.gov to efficiently search for suitable treatments for COVID-19 patients with various pre-existing conditions. AI4CoV was developed in the beginning of the pandemic when no treatment was known to be effective. AI4CoV helped by offering recommendations as to which treatment options were safe and suitable for each patient. AI4CoV can save time for healthcare teams to process thousands of clinical trial criteria. If widely deployed, AI4CoV can change clinical practices by allowing healthcare teams to consider a broad range of treatment options while avoiding those potentially harmful to patients, which may eventually lead to major improvement of public health. This is particularly useful when efficacy data of candidate treatments are not available at an early stage of a pandemic.

AI4CoV provides automated search for suitable clinical trials for a large number of patients during the pandemic to match the rapid pace of the growth of new treatment options registered for clinical trials. Since the eligibility criteria of clinical trials registered in ClinicalTrials.gov are given in itemized English text, Artificial Intelligence (AI) is necessary to quickly parse through the criteria in order to determine if a patient is eligible. For example, AI4CoV must comprehend that patients with “severe liver diseases,” among other exclusion criteria, need to be excluded from using drugs such as Remdesivir (NCT04252664, **Figure 1**). AI4CoV performs Natural Language Processing (NLP) to accomplish this task. AI4CoV’s NLP capability also allows it to read patient records in either structured data formats or unstructured clinical narrative text to extract patients’ clinical manifestations to determine if they are eligible for each criterion of a clinical trial. As such, AI4CoV can accommodate patient records in different structures and formats, a technical challenge known to hinder interoperability of electronic medical records.^9^

**Figure 1.**
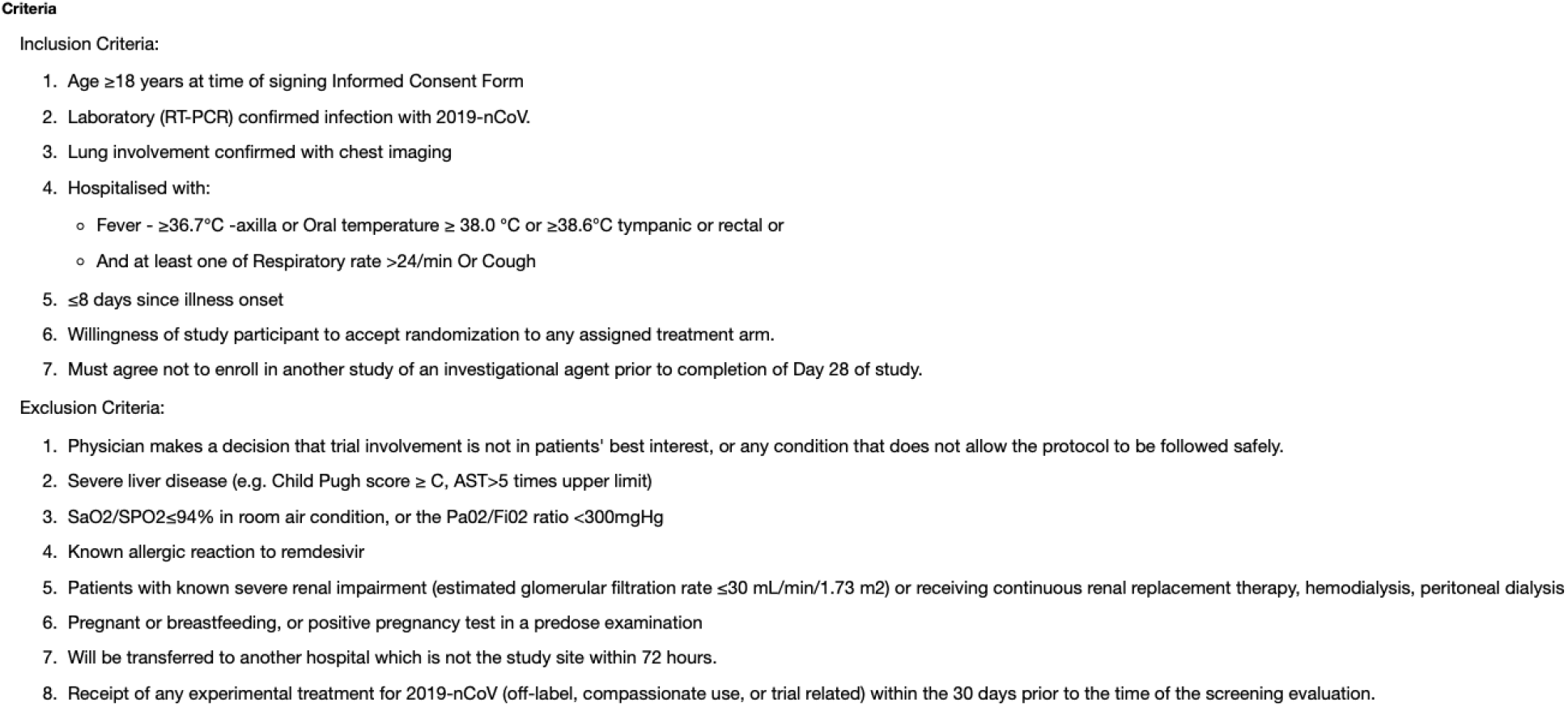
An example of clinical trial criteria listed in the ClinicalTrials.gov website (NCT04252664, Remdesivir). This example shows the complexity of the criteria. It also shows why it is necessary to use AI Natural Language Processing to determine patient-trial suitability if we need to deal with a large number of patient-trial pairs automatically.

AI4CoV can also be used to match eligible patients to clinical trials and facilitate patient recruitment. The methods for automatic matching have previously been studied intensively. Comprehensive reviews are available.^10,11^ Many studies attempted to simplify the task by classifying patients into common phenotypes.^12^ Ni et al.^13^ reported that their system, which did not apply any AI or machine learning algorithms, yielded 35.7% precision compared to the 83.22% by AI4CoV. Recently, the 2019 N2C2 challenge of medical informatics included a task on patient matching for clinical trials.^14^ The winners in that task utilized methods based on keyword/pattern matching, while Zhang et al.^15^ reported a deep learning approach that matches patients and trials for a broad range of diseases. AI4CoV is innovative and more effective than the aforementioned approaches since it takes advantage of both deep learning and pattern matching – deep learning helps by capturing semantically similar medical concepts (*e*.*g*., “arrythmia” and “QT prolongation”) while pattern matching allows for precise reasoning of quantitative clinical characteristics, *e*.*g*., “PaO2 ≥ 300” (partial pressure of oxygen in the arterial blood) and “SpO2 lower than 93%” (peripheral oxygen saturation). Determining if a patient satisfies these inequalities is clinically crucial and must be performed precisely.

Our performance evaluation shows that AI4CoV correctly predicts different drugs as suitable treatment options based on patients’ underlying conditions. Remarkably, in our retrospective review, AI4CoV predicted that three mild-to-moderate patients in our test cohort were safe to take Hydroxychloroquine while another patient would not have been due to having cardiac arrhythmia. all four patients were prescribed Hydroxychloroquine during their hospital stay. The aforementioned three patients were released from the hospital with no follow-up issues reported, while the patient predicted by AI4CoV as unsuitable to take Hydroxychloroquine was readmitted to the hospital within 30 days. If AI4CoV had been used, the healthcare team would have been alerted about the risk of Hydroxychloroquine for the patient.

For severe patients in our cohort, defined as those who were sent in an Intensive Care Unit, intubated, or subsequently died, AI4CoV predicted that generally have less treatment options suitable for them than the mild and moderate patients. Drugs such as Dexamethasone and Lopinavir-Ritonavir+Arbidol are unsuitable for all of our severe patients. While Hydroxychloroquine is suitable to only one of our severe patients. AI4CoV found that Interleukin-1 inhibitors (Anakinra) is a suitable option for half of our severe patients. Anakinra was suggested as a promising treatment of severe patients by several recent studies.^7,8^ AI4CoV also found that Ruxolitinib and Convalescent Plasma Transfer are suitable options for more than half of our severe patients. All other prominent drugs considered in our retrospective review have less than half of the severe patients with a suitable score.

## Methods

### Study Population

Our study cohort consists of 52 patients from two sites: 28 from the Taizhou Hospital, Taizhou, Zhejiang, China and 24 from the Lausanne University Hospital, Lausanne, Switzerland.

The Taizhou Hospital Institutional Review Board approved our study, in which the Strengthening the Reporting of Observational Studies in Epidemiology (STROBE) reporting guideline was followed. The patient data from the Lausanne University Hospital are available for download in the public domain^16^. The data were de-identified.

**Table 1** depicts the demographic and clinical characteristics of our study cohort. The 28 adult patients were admitted to the Taizhou Hospital with PCR-confirmed or clinically diagnosed COVID-19 between 20^th^ January and 27^th^ April 2020. We classified them by their COVID-19 status as mild or moderate using their PaO2 level, where 200 mmHg or above is mild and 100-200 mmHg is moderate. None of the Taizhou patients have the PaO2 level below 100 mmHg.

**Table 1.** Demographic and Clinical Characteristics of the Patient Cohort.

|  | Taizhou |  |  | Lausanne |
| --- | --- | --- | --- | --- |
| Condition | All (n=28) | Mild (n=22) | Moderate (n=6) | Severe (n=24) |
| Age(years) | 55.39±13.85(33,79) | 53.95±13.14(34,79) | 60.66±16.40(33,77) | 68.33±10.16(48,85) |
| Female | 12(42.85%) | 10(45.45%) | 2(33.33%) | 9(37.50%) |
| COVID-19 positive | 28(100%) | 22(100%) | 6(100%) | 24(100%) |
| Pneumonia | 14(50%) | 11(50%) | 3(50%) | No data |
| SpO2 | 92.67%±2.11(88,94) | 93.31%±1.32(89,94) | 90.33%±2.87(88,94) | 91.33%±6.18(73,97) |
| PaO2 (mmHg) | 241.11±62.37 (104,301) | 267.04±36.56 (212,301) | 146.00±39.73 (104,193) | No data |
| Fever | 24(85.71%) | 18(81.81%) | 6(100%) | 17(70%) |
| Cough | 27(96.43%) | 21(95.45%) | 6(100%) | 18(75%) |
| Sputum production | 22(78.57%) | 16(72.72%) | 6(100%) | 1(4.16%) |
| Sore-throat | 10(35.71%) | 8(36.36%) | 2(33%) | No data |
| Nasal discharge | 0(0%) | 0(0%) | 0(0%) | 3(12.5%) |
| Myalgia | 10(35.71%) | 9(40.9%) | 1(16%) | No data |
| Headache | 6(21.43%) | 4(18.18%) | 2(33%) | No data |
| Chest tightness | 24(85.71%) | 18(81.81%) | 6(100%) | 2(8.33%) |
| Dyspnea | 12(42.85%) | 8(36.36%) | 4(66%) | 19(79.16%) |
| Fatigue | 21(75%) | 15(68.18%) | 6(100%) | No data |
| Coronary artery disease | 8(28.57%) | 7(31.81%) | 1(16%) | No data |
| ALT > 5 | 1(3.57%) | 1(4.54%) | 0(0%) | 1(4.16%) |
| AST > 5 | 1(3.57%) | 1(4.54%) | 0(0%) | 1(4.16%) |
| High Blood Pressure | 0(0%) | 0(0%) | 0(0%) | 12(50%) |
| HBV positive | 2(7.14%) | 2(9.09%) | 0(0%) | No data |
| Autoimmune diseases | 1(3.57%) | 1(4.54%) | 0(0%) | 1(4.16%) |
| Bacterial Pneumonia | 1(3.57%) | 0(0%) | 1(16%) | No data |
| Arrhythmias | 1(3.57%) | 0(0%) | 1(16%) | No data |
| Warfarin tab | 1(3.57%) | 0(0%) | 1(16%) | No data |
| Value rep surgery | 1(3.57%) | 0(0%) | 1(16%) | No data |
| Diabetes | 3(10.71%) | 1(4.54%) | 2(33%) | 3(12.5%) |
| Fat Liver | 1(3.57%) | 1(4.54%) | 0(0%) | 1(4.16%) |
| Respiratory distress | 1(3.57%) | 0(0%) | 1(16%) | 4(16%) |
| Cancer | 0(0%) | 0(0%) | 0(0%) | 2(8.32%) |
| Entecavir<br>tab | 1(3.57%) | 1(4.54%) | 0(0%) | No data |
| IOT, ICU,<br>Death | 0(0%) | 0(0%) | 0(0%) | 24(100%) |
| Hospitalized | 28(100%) | 22(100%) | 6(100%) | 24(100%) |
| Symptom<br>max days | 15.35±7.31(2,31) | 13.72±6.64(2,27) | 21.33±7.00(12,31) | 8.16±4.94(2,21) |

A list of all notable patient attributes, including the COVID-19 classifications of mild, moderate, or severe. We classified all Lausanne University Hospital patients as severe because they were either deceased, in the ICU, or under intubation. There is a diverse distribution of Age in our Taizhou cohort.

To test AI4CoV for severe patients, 24 severe patients were selected from 80 patients from the Lausanne data set. Severe patients are those who, within seven days after they were admitted to the emergency department, were sent in an intensive care unit, intubated, or dead, the same criteria as defined in the original study where these patients were recruited.^17^ In comparison, none of the Taizhou patients satisfy these criteria. The demographic and clinical characteristics of these 24 severe patients are also given in **Table 1**.

We then selected 32 commonly considered clinical characteristics elements (**Table 1**) in COVID-19 studies,^18-20^ and then extracted these elements from the electronic medical record database of Taizhou patients and the data spreadsheet released by the Lausanne University Hospital for Lausanne patients, respectively. The extracted elements were used as the input patient records of AI4CoV to match with eligibility criteria of the COVID-19 clinical trials.

### Clinical Trials

We considered all clinical trials registered in ClinicalTrials.gov by 5^th^ June 2020, which totaled to 341,642 trials. Among them, ClinicalTrials.gov classified 1,982 as COVID-19 related. We continued to filter the trials by retaining trials with criteria of “Age 18-64, 65+” (N=1,964), “Interventional ONLY” (N=1,126), “Not yet recruiting OR recruiting OR enrolling by invitation OR active OR not recruiting OR completed” (N=1,103), excluding trials designed for healthcare workers (N=1,078), and excluding vaccines to finally obtain 1,062 unique trials. With 28 patients, we have a total of 29,736 patient-trial pairs to consider.

### Prominent Drugs

The study team selected 51 clinical trials targeting on 36 prominent drugs **(Table 2)** after reviewing publications and reports.^1,2,21,22,23,24^ Prominent drugs are treatment options of COVID-19 that have been widely reported in literature and are more likely to be considered by healthcare teams than other options, and thus may be more informative for performance evaluation of AI4CoV. The trial selection was independent of what drugs the patients had actually been prescribed. Trials comparing multiple target treatments were excluded. These trials were used in the evaluation of the predictive performance of AI4CoV.

**Table 2.**
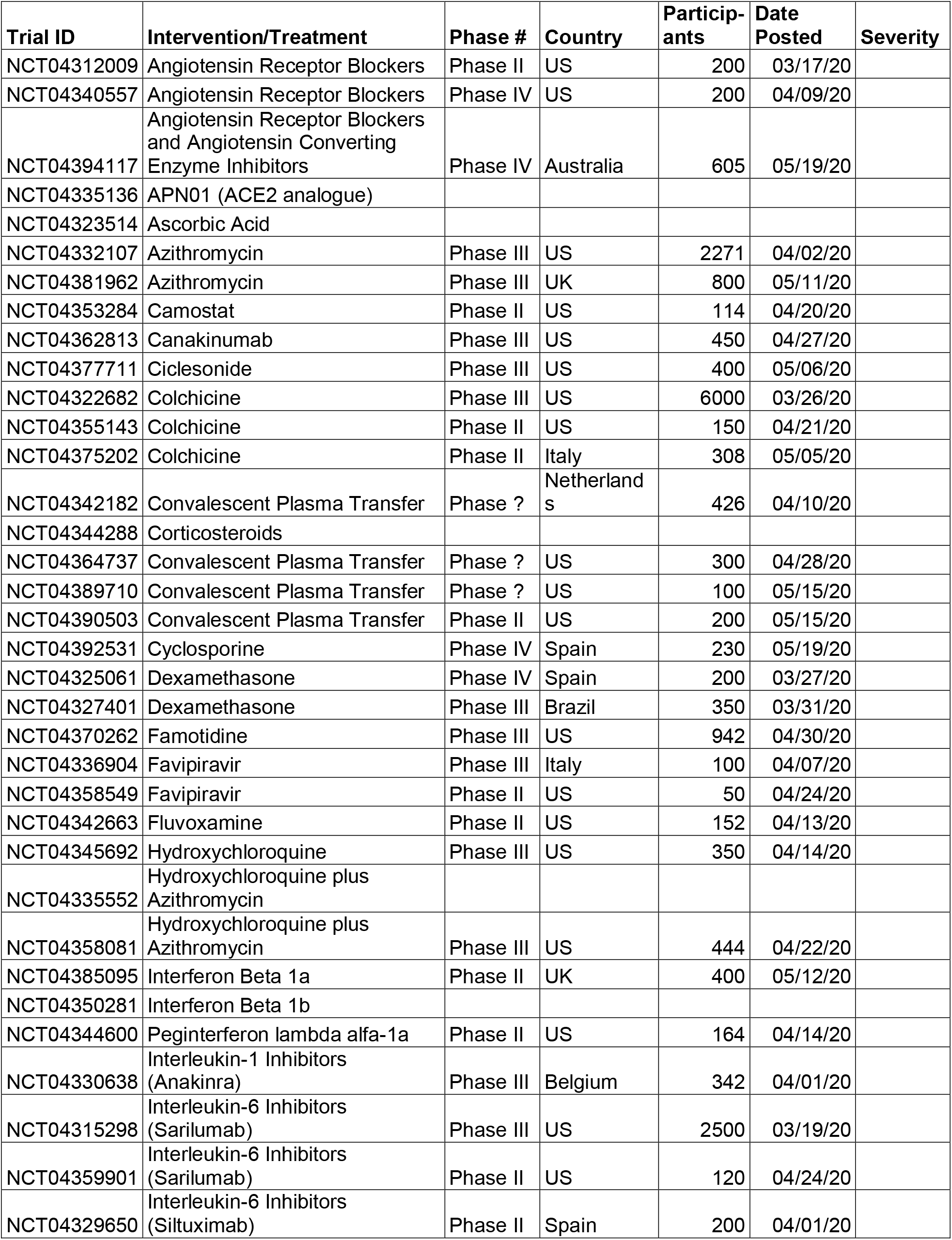
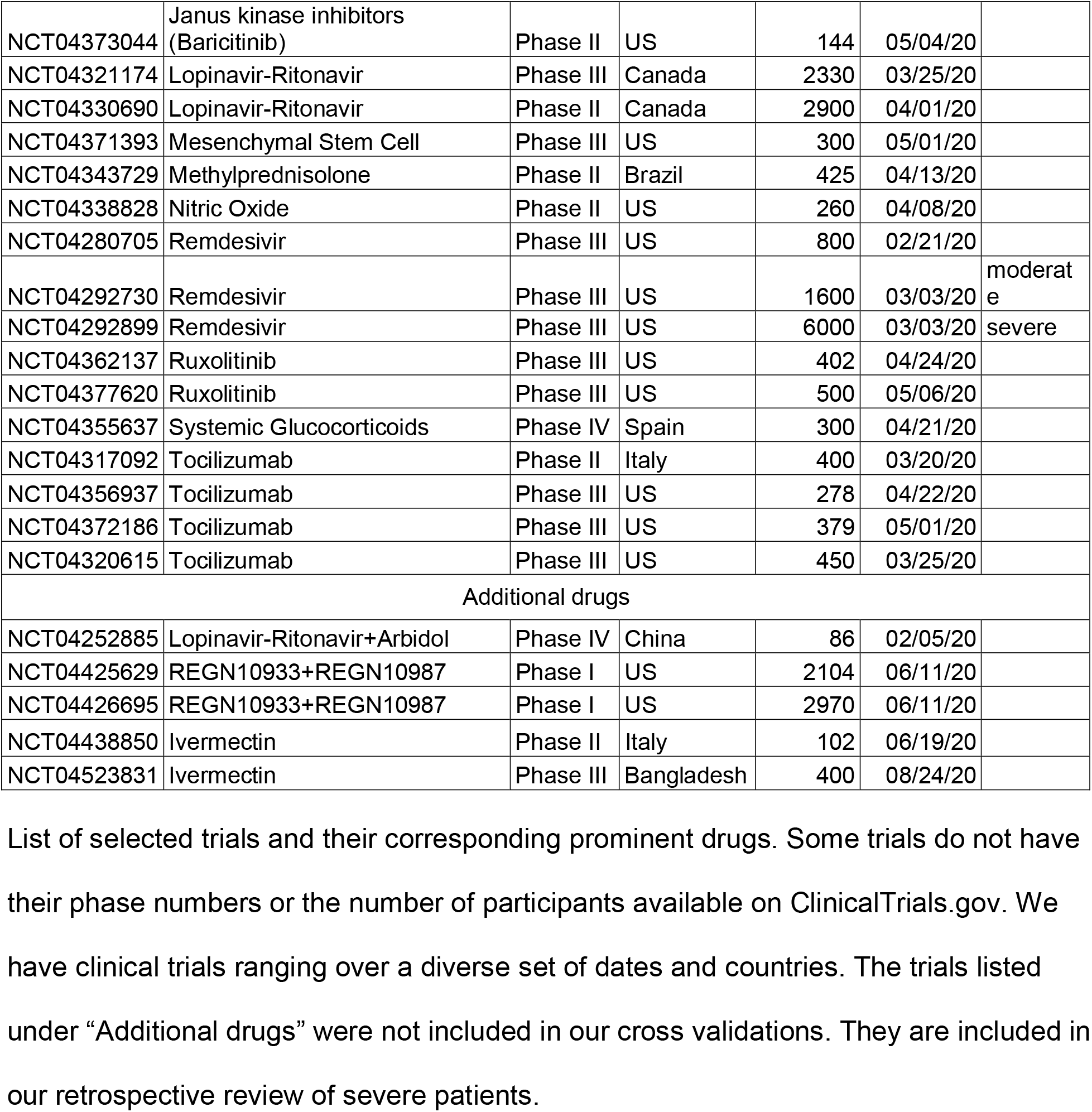
List of Clinical Trials of Prominent Drugs by NCT Number.

We considered the clinical trials of three additional drugs: REGN10933+REGN10987, Lopinavir-Ritonavir+Arbidol, and Ivermectin, in our retrospective review in addition to the aforementioned 36 prominent drugs because they were widely reported as emerging treatment options of COVID-19 after we completed our performance evaluation study. It is interesting to see if AI4CoV predicts them as suitable for the patients in our cohort. The selected clinical trials are also given in **(Table 2)**.

### Performance Evaluation

The study team sampled and reviewed 3,156 patient-trial pairs from the 29,736 total pairs and assigned each pair a label of either “suitable” or “unsuitable” to create the ground truth. A patient-trial pair is *suitable* if within the pair, the patient satisfies the trial’s inclusion criteria and does not satisfy any exclusion criteria; otherwise it is labeled as *unsuitable*. The records of the 28 Taizhou patients were used to create the 3,156 ground-truth patient-trial pairs. The records of the 24 Lausanne severe patients were reserved for validation.

We developed and compared the performance of six versions of AI4CoV in this study. To evaluate the performance of each version, a 10-fold cross-validation and a 7-fold cross-validation were used. In the 10-fold cross-validation, 3,156 pairs were randomly divided into 10 folds for train-test splitting. The evaluation assumed that each patient-trial pair is independent.

In the 7-fold cross-validation, 28 Taizhou patients were divided randomly into seven folds. As such, no patient appeared in both training and test sets, which simulates real situations where the system is trained on known patients and is used to select trials and drugs for new, unseen patients. In this evaluation, the seven patients in the test set in each fold were only paired with the 51 trials of the prominent drugs. That summed to a total of 1,428 pairs of patient-trials being tested in the seven folds. The training set in each fold included all available patient-trial pairs involving the other 24 Taizhou patients.

We assessed each version of AI4CoV by comparing the predicted scores of suitability to the ground truth labels. We calculated sensitivity, specificity, precision, negative predictive value, AUROC (area under the receiver operating characteristic curve), and F1-score to measure AI4CoV’s performance. Since we wanted to assess the discriminative ability (*i*.*e*., how well AI4CoV predicts binary patient-trial suitability), goodness-of-fit statistics for calibration were not considered.^25^

### The AI4CoV Algorithm

AI4CoV reads patient records and clinical trial criteria from ClinicalTrials.gov as the input and assigns a score indicating the strength of the trial’s suitability to the patient for each patient-trial pair **(Figure 2)**.

**Figure 2.**
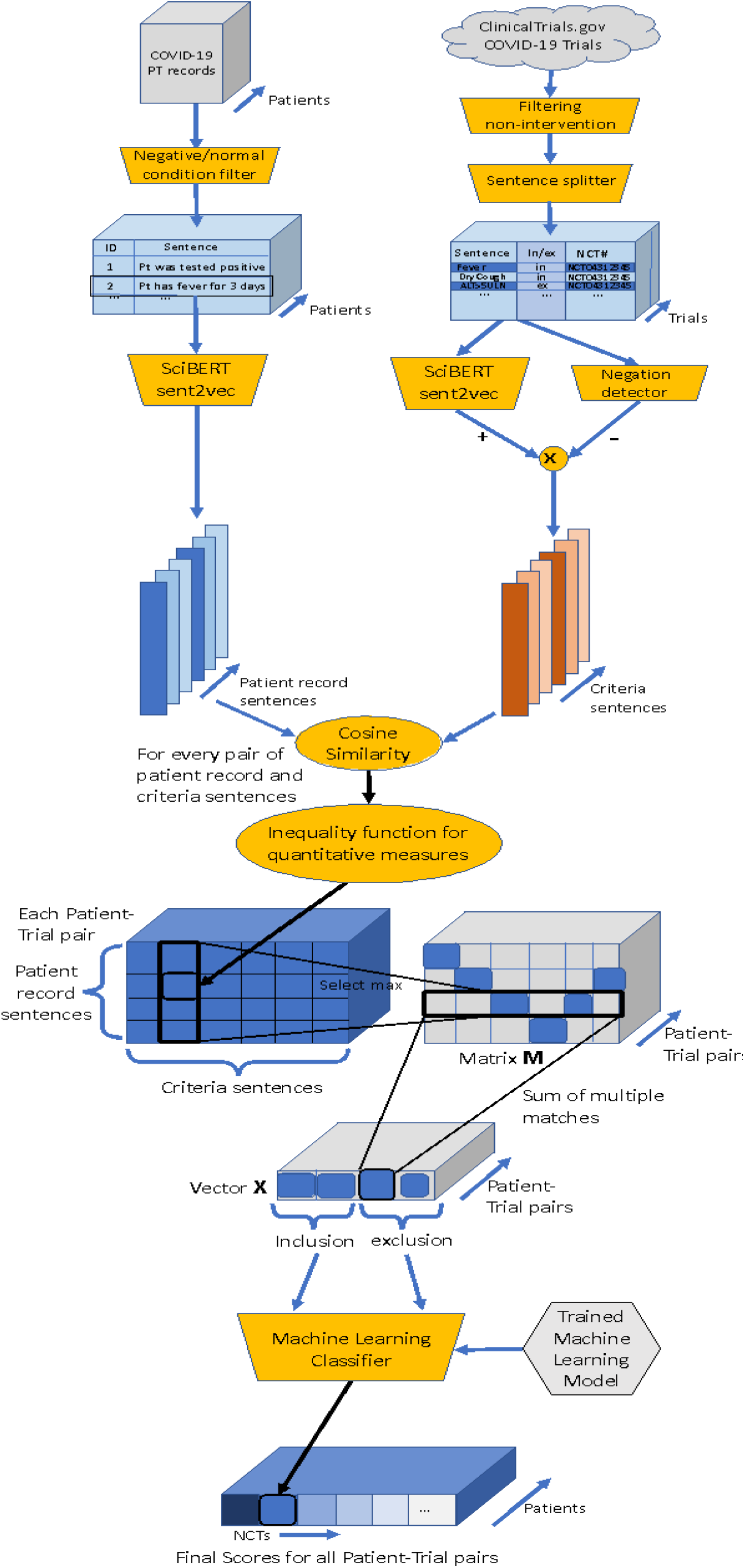
Workflow of AI4CoV. Details are described in “The AI4CoV Algorithm” section in “Method.”

For every clinical trial under consideration, AI4CoV applies a preprocessing step to split its eligibility criteria into a text file with each line containing three columns: a criteria sentence, “in” or “ex” depending on whether the criteria is an inclusion or exclusion, and the NCT number of the trial assigned by ClinicalTrials.gov. Each criteria sentence must be “atomic” in the sense that the sentence describes a criterion defined on a clinical manifestation and cannot be further divided. For example, “patients with malignant tumors” is atomic but “untreated active hepatitis or HIV-positive patients” is not and should be further divided into two sentences for the manifestations of “hepatitis” and “HIV-positive.”

After creating these text files, AI4CoV uses the BioSent2vec function to convert each atomic criterion into a vector of 700 dimensions to represent its semantics.^26^ BioSent2vec is a deep neural network language model trained by a large corpus of biomedical research papers to convert a sentence into a vector such that sentences of similar semantics are close to each other in the vector space. Next, a NegEx function multiplies 1 or −1 to the vector depending on whether the atomic criterion expresses a negation (for example, “no fever”).^27^

Each patient record inputted contains demographics and clinical manifestations (**Table 1**). Again, a preprocessing step converts each patient record into a text file where each line contains a “patient record sentence.” A patient record sentence must be atomic as defined for the criteria sentences above. Patient record sentences that describe normal manifestations as negated statements are discarded. For example, “fever for 3 days” is retained while “no history of hypertension” is not. BioSent2vec is then applied to convert each patient characteristic into a vector of 700 dimensions.

After obtaining 700-dimension vectors for patients and clinical trials, AI4CoV calculates the cosine similarity of each patient record sentence vector and clinical trial criteria sentence vector pair to estimate how semantically similar these sentences are. This step creates a matrix M of cosine similarity scores for each patient-trial pair.

As an optional step to mitigate the limitation of cosine similarity, the inequality function deals with quantitative clinical characteristics such as “Age,” “SpO2,” and “PaO2.”. If both patient record and criterion sentence share the same keywords of these measures, the inequality function finds the nearest number and inequality sign, such as ≥, <, “less,” etc., after the keyword and compares the patient record’s value and the criteria value to determine if the criterion is satisfied. If so, the cosine similarity score is retained in the matrix M; otherwise, a negative number is assigned. In our implementation, “-5” was used.

After all patient sentence and criteria sentence pairs have a score, AI4CoV calculates the maximum cosine similarity score for each criterion and erases other scores in M. If the criterion has a score of “-5” then the “-5” will be retained instead of the maximum score. From the resulting matrix, AI4CoV then creates an array X of 64 elements by number of trial-patient pairs, where 32 elements for common COVID-19 clinical characteristics represented inclusion criteria and the other 32 represented exclusion criteria. Each element contains the retained score for each row of M. The score estimates the matching strength of a patient record clinical characteristic to an inclusion or exclusion criterion. When no match is found, the element for that clinical characteristic is zero. If multiple criterion sentences from the same trial have a score corresponding to the same patient record sentence, then AI4CoV sums all scores and places the result in X (**Figure 2**).

From array X, we developed six versions of AI4CoV and compared their performances

⍰ Baseline: Sum of the cosine similarity scores of the inclusion criteria in X minus the sum of those of the exclusion criteria as the score of suitability.
⍰ Baseline with inequality: Same as Baseline but the inequality function is used in addition to cosine similarity.
⍰ NN: A multiple-layer-perceptron neural network with a hidden layer of 120 nodes is trained with labeled suitability scores to classify an array X as suitable or unsuitable.
⍰ NN with inequality: Same as NN but the inequality function is used as well.
⍰ XGBoost: An XGBoost classifier^28^ is trained to classify X.
⍰ XGBoost with inequality: Same as XGBoost but the inequality function is used as well.

Other machine learning models that we tested include Logistic Regression, Gradient Boosting Classifier, and C-Support Vector Classification. But none were found to be as effective as the two that are reported.

### Retrospective Review

We performed a retrospective review of the medication used on the Taizhou patients in our cohort and their outcomes within 60 days of hospital admission. The outcome metrics considered were based on observational studies.^29,30^ Predicted suitability results by the best performing AI4CoV version were compared to the medication used and resulting outcomes of the patients.

We also applied the best performing AI4CoV version to patient-trial pairs of the 24 Lausanne severe patients and the clinical trials in **Table 2** and compared the distributions of the predicted scores by AI4CoV among these patients and with the 28 Taizhou patients.

## Results

### Performance of AI4CoV

**Table 3** summarizes all of the versions that we evaluated by the 7-fold and 10-fold cross-validation. **Figure 3** shows the graphs of ROC curves. The best performing version was XGBoost with the inequality function, achieving 92.76% AUROC and 87.40% F1-score in the 7-fold cross validation. It also performed the best under 10-fold cross validation, while NN with the inequality function finished a close second, and the two Baselines finished last. Versions with the inequality function achieved higher AUROC scores than those without.

**Table 3.** Performance Metrics Comparison of the Six Versions of AI4CoV.

| Version | AUROC | Sensitivity | Specificity | Precision | Negative Predictive Value | Accuracy | F1-Score |
| --- | --- | --- | --- | --- | --- | --- | --- |
| 7-Fold Cross-Validation |  |  |  |  |  |  |  |
| Baseline | 75.52% | 60.05% | 58.53% | 69.36% | 48.37% | 59.45% | 64.37% |
| Baseline + ineq | 76.87% | 54.99% | 68.76% | 73.35% | 49.42% | 60.36% | 62.86% |
| NN | 89.62% | 88.29% | 64.63% | 79.61% | 77.92% | 79.06% | 83.72% |
| NN + inequality | 90.03% | 89.44% | 65.17% | 80.06% | 79.78% | 79.97% | 84.49% |
| XGBoost | 92.39% | 91.04% | 70.38% | 82.78% | 83.40% | 82.98% | 86.71% |
| XGBoost + ineq | **92.76%** | **92.42%** | **70.20%** | **82.90%** | **85.56%** | **83.75%** | **87.40%** |
| 10-Fold Cross-Validation |  |  |  |  |  |  |  |
| Baseline | 76.70% | 65.77% | 50.55% | 66.51% | 49.73% | 59.66% | 66.13% |
| Baseline + ineq | 77.55% | 60.26% | 57.66% | 68.00% | 49.29% | 59.22% | 63.90% |
| NN | 87.14% | 82.06% | 70.14% | 80.40% | 72.37% | 77.28% | 81.23% |
| NN + inequality | 87.46% | 82.17% | 70.93% | 80.84% | 72.71% | 77.66% | 81.50% |
| XGBoost | 89.75% | 87.72% | 72.20% | 82.49% | 79.76% | 81.50% | 85.03% |
| XGBoost + ineq | **90.25%** | **88.68%** | **73.30%** | **83.22%** | **81.26%** | **82.51%** | **85.86%** |

7-Fold cross-validation results for all selected trials: Baseline vs.
Fold cross-validation results for prominent drug trials: Baseline vs. Baseline+inequality
vs. NN vs. NN+inequality vs. XGBoost vs. XGBoost+inequality. Numbers in bold fonts
represent the best performance by the compared versions.

**Figure 3.**
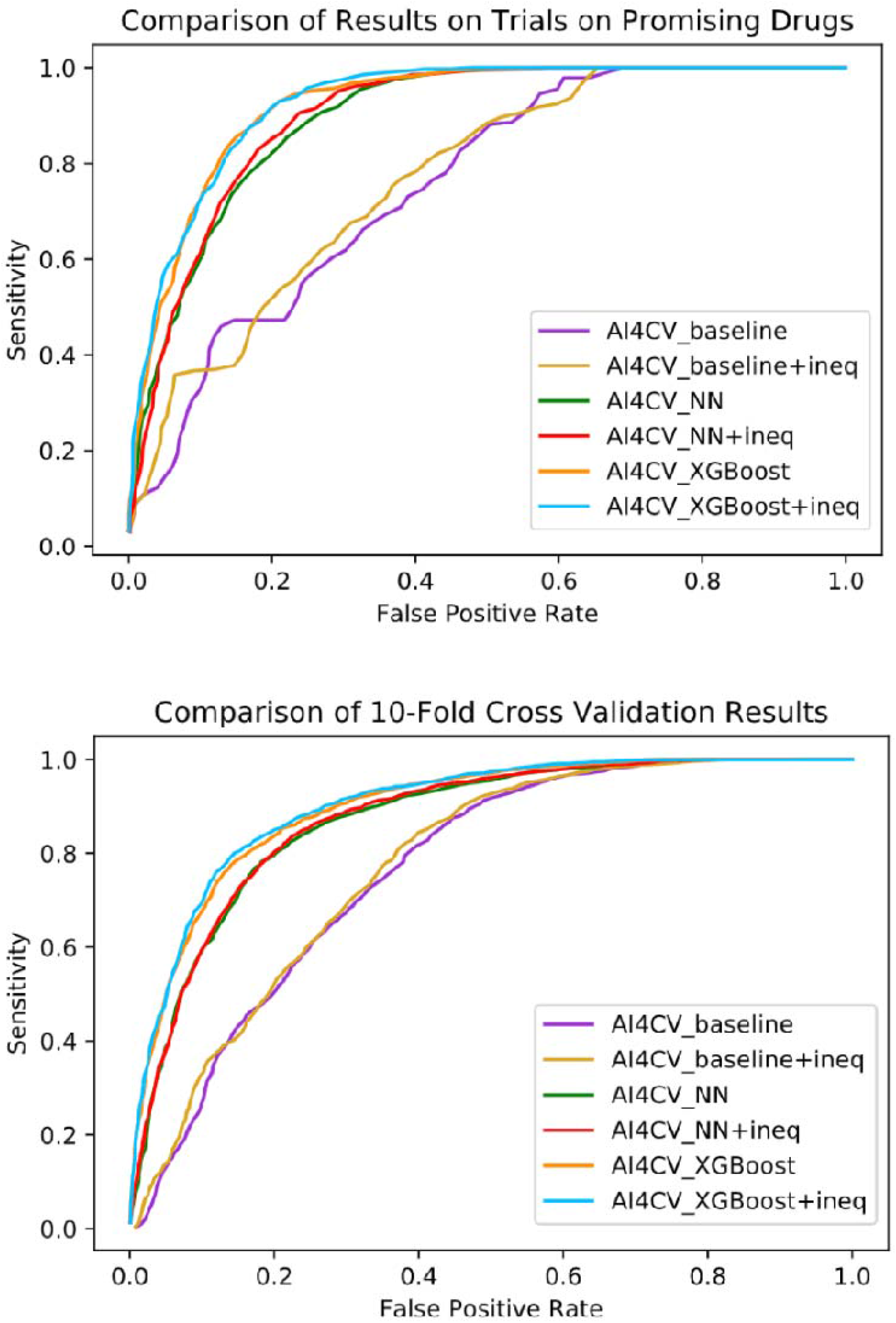
ROC Curves. (Top) ROC curve for 7-fold cross-validation, prominent drug trials: Baseline vs. Baseline+inequality vs. NN vs. NN+inequality vs. XGBoost vs. XGBoost+inequality. (Bottom) ROC curve for 10-fold cross-validation, all patient-trial pairs with human labels: Baseline vs. Baseline+inequality vs. NN vs. NN+inequality vs. XGBoost vs. XGBoost+inequality. The ROC curves show that patient-trial pairs that have high scores by AI4CoV are indeed more likely to be suitable than those that have low scores.

### Results of Retrospective Review

All of the 28 patients from Taizhou had an incidence of pneumonia within 60 days of hospital admission but were all eventually discharged. Patients 12 and 15 also had an incidence of acute respiratory failure. Patient 21 had arrhythmia before and after treatment. Two patients were readmitted to the hospital within 30 days and were eventually discharged again.

In total, the 28 patients took about fifty drugs combined, including traditional Chinese medicine, during their stay in the hospital. Most medications prescribed were for symptomatic treatment and are not discussed below. We compared the best performing AI4CoV (XGBoost with inequality) predicted suitability scores for the pairs of the patients and the clinical trials that tested the most common drugs that the patients were prescribed **(Table 4)**, specifically, Interferon alpha-2b, Methylprednisolone,

**Table 4.** Retrospective Review of Patient Outcomes and Medications.

| Patient | Outcomes within 60 days? | COVID-19 Medication | Suitability? (+ or -) | Average Suitability Score |
| --- | --- | --- | --- | --- |
| 1 | Incidence Pneumonia | Interferon alpha-2b | -, -, + | -1.26 |
|  |  | Methylprednisolone | + | 1.96 |
|  |  | Lopinavir-Ritonavir | -, + | -0.37 |
| 2 | Incidence Pneumonia | Interferon alpha-2b | -, -, + | -1.02 |
|  |  | Methylprednisolone | + | 0.75 |
|  |  | Lopinavir-Ritonavir | -, + | -0.03 |
|  |  | Immunoglobulin | +, +, +, +, +, - | 1.98 |
|  |  | Azithromycin | -, +, +, + | 1.52 |
|  |  | Hydroxychloroquine | +, +, + | 2.97 |
| 3 | Incidence Pneumonia | Interferon alpha-2b | -, -, + | -0.74 |
|  |  | Methylprednisolone | + | 2.36 |
|  |  | Lopinavir-Ritonavir | -, + | -0.18 |
|  |  | Azithromycin | +, +, +, + | 2 |
| 4 | Incidence Pneumonia | Interferon alpha-2b | -, -, - | -2.04 |
|  |  | Methylprednisolone | - | -2.51 |
|  |  | Lopinavir-Ritonavir | -, + | -0.2 |
| 5 | Incidence Pneumonia | Interferon alpha-2b | -, -, + | -2.15 |
|  |  | Methylprednisolone | + | 1.58 |
|  |  | Immunoglobulin | +, +, +, +, +, + | 1.36 |
|  |  | Lopinavir-Ritonavir | -, + | -1.26 |
|  |  | Hydroxychloroquine | +, +, - | 2.7 |
| 7 | Incidence Pneumonia | Interferon alpha-2b | -, -, + | -0.74 |
|  |  | Methylprednisolone | + | 1.03 |
|  |  | Immunoglobulin | +, +, -, +, +, + | 1.5 |
|  |  | Lopinavir-Ritonavir | -, + | 0.89 |
| 8 | Incidence Pneumonia | Interferon alpha-2b | -, -, + | -1.71 |
|  |  | Methylprednisolone | + | 0.59 |
|  |  | Immunoglobulin | +, +, -, +, +, + | 1.81 |
|  |  | Lopinavir-Ritonavir | -, + | -0.62 |
| 9 | Incidence Pneumonia | Interferon alpha-2b | -, -, + | -1.19 |
|  |  | Methylprednisolone | + | 1.48 |
|  |  | Immunoglobulin | +, +, +, +, -, - | 1.3 |
|  |  | Lopinavir-Ritonavir | -, + | 0.12 |
| 10 | Incidence Pneumonia | Interferon alpha-2b | -, -, - | -2.3 |
|  |  | Methylprednisolone | - | -0.09 |
|  |  | Lopinavir-Ritonavir | -,+ | -2.09 |
| 11 | Incidence Pneumonia | Interferon alpha-2b | -, -, + | -0.7 |
|  |  | Methylprednisolone | + | 2.56 |
|  |  | Immunoglobulin | +, +, +, +, +, + | 2 |
|  |  | Lopinavir-Ritonavir | +, + | 0.45 |
| 12 | Incidence Acute Respiratory Failure and Incidence Pneumonia | Interferon alpha-2b | -, -, + | -2.15 |
|  |  | Methylprednisolone | + | 2.02 |
|  |  | Immunoglobulin | +, +, +, +, +, + | 1.7 |
|  |  | Lopinavir-Ritonavir | +, + | 1.3 |
| 13 | Incidence Pneumonia | Interferon alpha-2b | -, -, + | -1.6 |
|  |  | Methylprednisolone | + | 0.62 |
|  |  | Lopinavir-Ritonavir | -, + | -0.55 |
|  |  | Immunoglobulin | +, +, -, +, +, + | 1.18 |
| 14 | Incidence Pneumonia | Interferon alpha-2b | -, -, + | -1.59 |
|  |  | Methylprednisolone | + | 1.48 |
|  |  | Lopinavir-Ritonavir | -, + | -1.09 |
| 15 | Incidence Acute Respiratory Failure and incidence Pneumonia | Interferon alpha-2b | -, -, + | -1.67 |
|  |  | Methylprednisolone | - | -3.81 |
|  |  | Lopinavir-Ritonavir | -, + | -0.41 |
|  |  | Immunoglobulin | -, +, +, +, -, + | 0.27 |
| 16 | Incidence Pneumonia | Interferon alpha-2b | -, -, + | -1.65 |
|  |  | Methylprednisolone | + | 1.45 |
|  |  | Immunoglobulin | +, +, +, +, +, + | 1.86 |
|  |  | Lopinavir-Ritonavir | -, + | -0.52 |
| 17 | Incidence Pneumonia | Interferon alpha-2b | -, -, + | -2.24 |
|  |  | Methylprednisolone | + | 0.16 |
|  |  | Immunoglobulin | +, +, -, +, -, - | 0.86 |
|  |  | Lopinavir-Ritonavir | -, + | -0.72 |
| 18 | Incidence Pneumonia | Interferon alpha-2b | -, -, + | -1.12 |
|  |  | Methylprednisolone | + | 1.61 |
|  |  | Lopinavir-Ritonavir | -, + | -0.71 |
| 19 | Incidence Pneumonia | Interferon alpha-2b | -, -, + | -1.34 |
|  |  | Methylprednisolone | + | 1.54 |
|  |  | Lopinavir-Ritonavir | +, + | 0.51 |
|  |  | Immunoglobulin | +, +, +, +, +, + | 1.17 |
| 20 | Incidence Pneumonia | Interferon alpha-2b | -, -, + | -1.43 |
|  |  | Methylprednisolone | + | 0.88 |
|  |  | Lopinavir-Ritonavir | +, + | 0.71 |
|  |  | Immunoglobulin | +, +, -, +, +, + | 1.57 |
| 21 | Prevalence Arrhythmia<br>Incidence Pneumonia | Interferon alpha-2b | -, -, + | -1.44 |
|  |  | Methylprednisolone | - | -3.78 |
|  |  | Lopinavir-Ritonavir | +, + | 2 |
|  |  | Immunoglobulin | +, +, -, +, +, + | -0.05 |
|  |  | Hydroxychloroquine | -, -, - | -1.77 |
| 22 | Incidence Pneumonia | Interferon alpha-2b | -, -, + | -1.2 |
|  |  | Methylprednisolone | + | 0.14 |
|  |  | Lopinavir-Ritonavir | +, + | 2.07 |
| 23 | Incidence Pneumonia | Interferon alpha-2b | -, -, + | -1.69 |
|  |  | Methylprednisolone | + | 0.14 |
|  |  | Lopinavir-Ritonavir | +, + | 1.1 |
|  |  | Immunoglobulin | +, +, -, +, +, + | 1.72 |
| 24 | Incidence Pneumonia | Interferon alpha-2b | -, -, + | -1.25 |
|  |  | Methylprednisolone | + | 0.28 |
|  |  | Lopinavir-Ritonavir | +, + | 3.27 |
|  |  | Immunoglobulin | +, +, -, +, +, + | 1.29 |
| 25 | Incidence Pneumonia | Interferon alpha-2b | -, -, + | -2.2 |
|  |  | Methylprednisolone | + | 0.88 |
|  |  | Lopinavir-Ritonavir | +, + | 0.12 |
|  |  | Immunoglobulin | +, +, +, +, +, + | 1.5 |
| 26 | Incidence Pneumonia | Interferon alpha-2b | -, -, + | -1.71 |
|  |  | Methylprednisolone | + | 2.42 |
|  |  | Lopinavir-Ritonavir | +, + | 1.14 |
|  |  | Immunoglobulin | +, +, -, +, +, + | 1.31 |
| 27 | Incidence Pneumonia | Interferon alpha-2b | -, -, + | -0.55 |
|  |  | Methylprednisolone | + | 1.25 |
|  |  | Lopinavir-Ritonavir | -, + | -0.2 |
|  |  | Immunoglobulin | +, +, -, +, -, + | 1.02 |
| 28 | Incidence Pneumonia | Interferon alpha-2b | -, -, + | -0.91 |
|  |  | Methylprednisolone | + | 1.03 |
|  |  | Lopinavir-Ritonavir | -, - | -0.83 |
|  |  | Immunoglobulin | +, +, -, +, +, + | 1.81 |
|  |  | Hydroxychloroquine | +, -, + | 0.6 |
| 29 | Incidence Pneumonia | Interferon alpha-2b | -, -, + | -1.9 |
|  |  | Methylprednisolone | + | 1.14 |
|  |  | Immunoglobulin | +, +, -, +, +, + | 1.45 |
|  |  | Lopinavir-Ritonavir | -, + | 0.74 |

Results of the retrospective review of the outcomes of the patients and the medications they took, compared with their AI4CoV predicted average suitability scores for each medication. The outcome of patient 6 is incomplete and was thus excluded from the study. Outcomes marked as “Incidence” signifies that the patient gained the condition after hospital admission, while “Prevalence” indicates that the patient had the condition before hospital admission. COVID-19 medications for each patient are ordered by the first date that they were prescribed to the patient. Each medication may have a different number of registered clinical trials. Column “Suitability? (+ or -)” shows whether the patient-clinical trial pair has a positive (suitable) or negative (unsuitable) score as predicted by AI4CoV. The ordering of plus and minus for each drug is consistent across all patients to allow for comparison of whether a trial yielded a negative or positive score for most patients. The last column shows the average score of the trials for each medication.

Immunoglobulin, Lopinavir-Ritonavir, Hydroxychloroquine, and Azithromycin. We note that for each drug there are multiple clinical trials registered and considered here. Overall, Methylprednisolone, Immunoglobulin, Hydroxychloroquine, and Azithromycin yielded suitable average scores while Interferon alpha-2b and Lopinavir-Ritonavir, two drugs that all 28 patients were prescribed, yielded unsuitable average scores.

Since none of the 28 patients from Taizhou are severe, we tested AI4CoV with the 24 severe patients from the Lausanne University Hospital. **Figure 4** shows the range of suitability scores for all Lausanne University Hospital severe patients (that were in ICU, intubated, or died within seven days) and the prominent drug trial pairs. The drug names are listed on the x-axis. Most of the scores are below zero, indicating that most of the prominent drugs were predicted as not suitable for the severe patients by AI4CoV. This is expected because clinical trials generally place more restrictions for patients with a variety of underlying conditions to minimize adversarial events and severe patients need to take additional precautions in their treatment selection. The scores from AI4CoV allow for estimation and comparison of how strict or lenient the eligibility criteria of the clinical trials can be.

**Figure 4.**
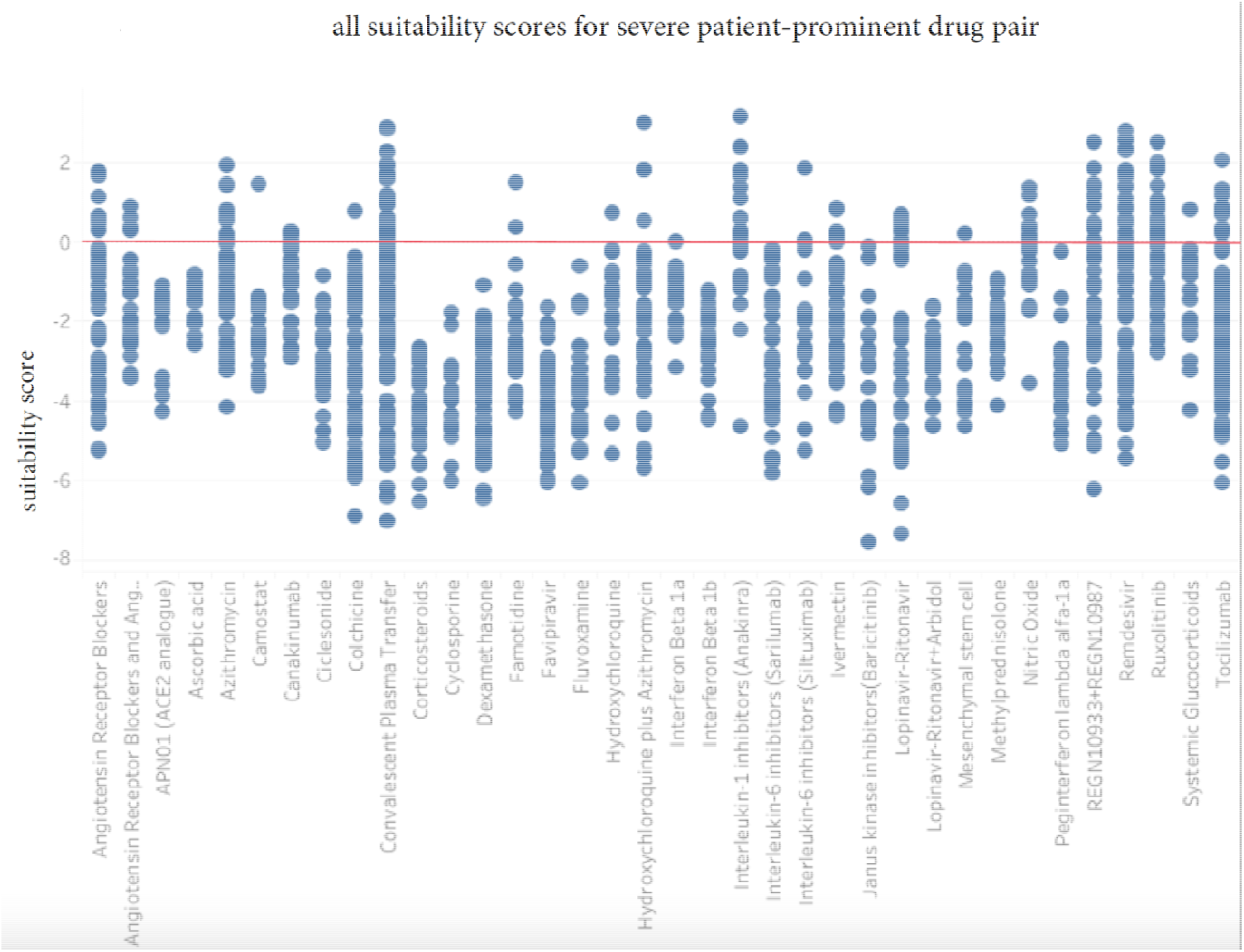
The range of suitability scores for all severe patients from Lausanne University Hospital-prominent drug trial pair. The drug names are listed on the x-axis. It shows that some drugs such as Convalescent Plasma Transfer and Anakinra have many suitable severe patients, while some drugs such as Dexamethasone and Hydroxychloroquine are unsuitable to almost all severe patients. Most drugs are only suitable to take for a few patients.

According to AI4CoV, severe patients generally have less treatment options than mild and moderate patients, because they have more drugs suitable for them than the severe patients **(Figure 5)**.

**Figure 5.**
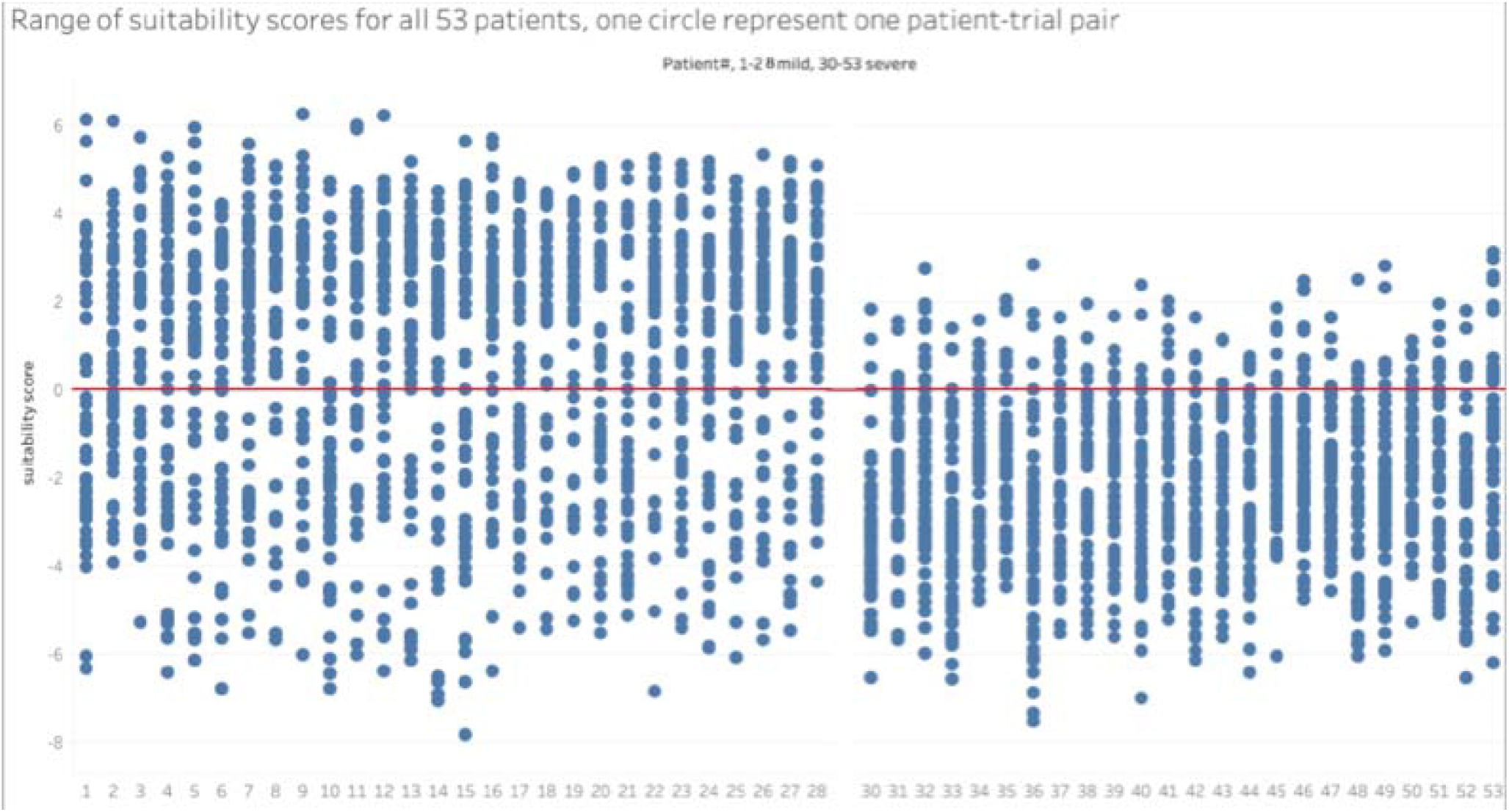
This graph shows that mild and moderate patients (1-29) have more treatment options than severe patients (30-53). All patients have some safe treatment options available.

Our results suggest that AI4CoV can help healthcare teams search for treatment options from clinical trials for COVID-19 patients. Healthcare teams for COVID-19 may reference **Table 5** to check the top criteria used by all interventional COVID-19 clinical trials as well as the top criteria used by the 51 clinical trials for the prominent drugs. A list of top criteria was manually compiled previously.^31^ AI4CoV makes it possible to maintain an up-to-date list automatically.

**Table 5.**
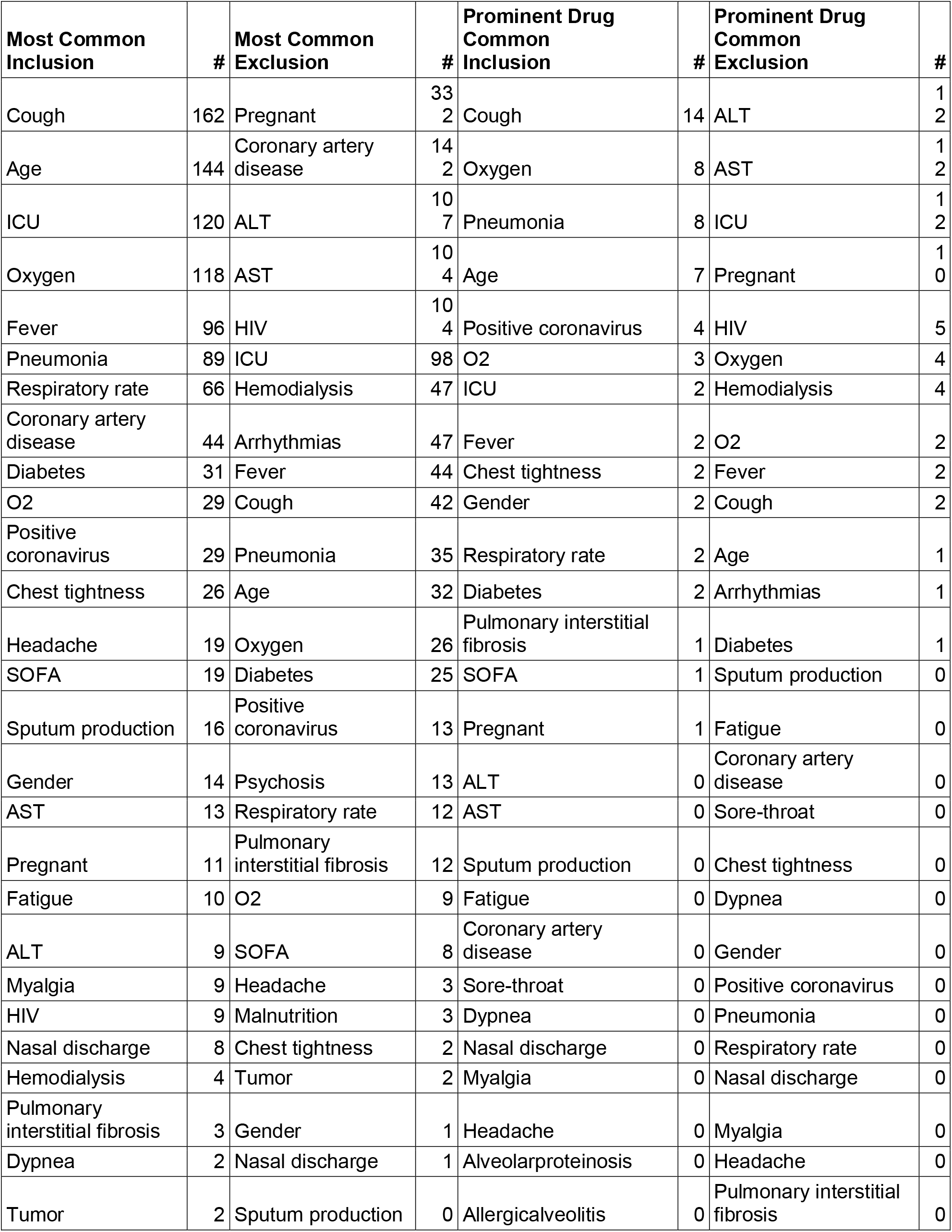
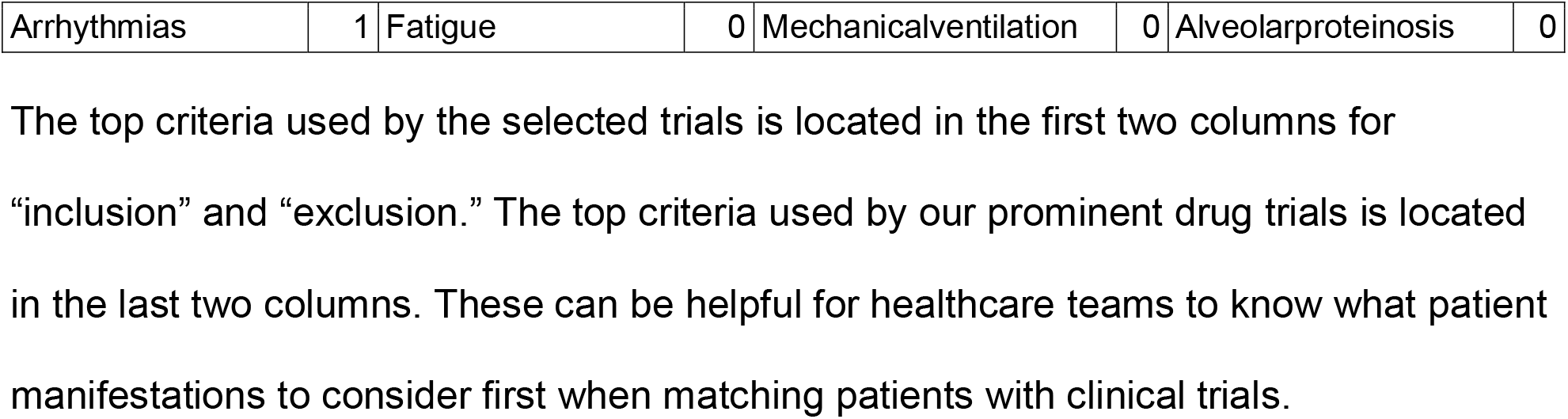
Most Frequent Criteria.

## Discussion

The best performing version of AI4CoV leverages three key AI techniques: (1) BioSent2vec,^26^ (2) rule-based inequality function, and (3) XGBoost^28^ machine learning classifier. BioSent2vec is a deep neural network language model that allows AI4CoV to correctly match semantically similar clinical manifestations of patients with eligibility criteria of clinical trials, while a rule-based pattern matching approach to precise modeling of inequalities that express quantitative clinical manifestations such as “PaO2 ≥ 300” is crucial because it is still challenging for state-of-the-art deep neural network language models to deal with numbers and inequalities without a large number of labeled training examples to fine-tune the models. We opted to a hybrid design to leverage the advantages of both approaches. XGBoost outperformed a neural network in our evaluation. This may be because decision trees that XGBoost learns can model the importance of clinical features more effectively than the weights that a neural network learns in AI4CoV’s criterion matching task. Overall, we show that by integrating these modules, AI4CoV achieves high performance with a hybrid approach.

In our retrospective review, all Taizhou mild and moderate patients in our study cohort took Methylprednisolone. 25 out of 28 patients have a suitable score to the clinical trial NCT04343729, which tests Methylprednisolone as an intervention to treat COVID-19. While most patients recovered successfully without readmitted to the hospital, patient 21 **(Table 1)** was given a negative suitability score for Methylprednisolone, which matched his readmittance to the hospital within 30 days. AI4CoV also gave patient 15, who had an incidence of acute respiratory failure, a negative score for Methylprednisolone. Meanwhile, most patients recovered successfully and were released from the hospital without complications. This is an example of AI4CoV correctly indicating that Methylprednisolone is a safe drug for most of the patient cohort, and correctly singling out specific patients for whom the drug would have been unsafe.

Patients 2, 5, 21, and 28 also took Hydroxychloroquine. Remarkably, patients 2, 5, and 28, all of whom AI4CoV gave average positive (suitable) scores, were discharged from the hospital with no further issues reported. In contrast, patient 21, for whom AI4CoV gave negative (unsuitable) scores for all three clinical trials on Hydroxychloroquine, was readmitted to the hospital within 30 days with persistent arrhythmia. From these cases, AI4CoV accurately predicted that it was unsafe for patient 21 to take Hydroxychloroquine while it was safe for the other three patients to consider Hydroxychloroquine as a treatment option. This demonstrates the effectiveness of using BioSent2vec^26^ within AI4CoV’s algorithm because although the exclusion criteria did not explicitly contain “Arrhythmia”, it did state phrases closely related to Arrhythmia, such as “QT prolongation” and “ECG abnormality”. BioSent2vec is a deep neural network language model specialized in the language of biomedical sciences. AI4CoV correctly identified these phrases and recognized that patient 21 with arrhythmia should be excluded from taking Hydroxychloroquine. We note that patient 21 took both Hydroxychloroquine and Methylprednisolone. Both were unsuitable for the patient and should have been excluded.

**Figure 4** shows that drugs such as Dexamethasone and Lopinavir-Ritonavir+Arbidol are unsuitable for all of our severe patients, while Hydroxychloroquine is suitable to only one of our severe patients. AI4CoV found that Interleukin-1 inhibitors (Anakinra) is a suitable option for half of our 24 severe patients, matching the suggestion of several recent studies. AI4CoV also found that Ruxolitinib, and Convalescent Plasma Transfer are suitable options for more than half of our 24 severe patients. All other prominent drugs were determined to give less than half of severe patients a suitable. This is due to the fact that trials utilizing Dexamethasone and Hydroxychloroquine contain stricter eligibility criteria, such as requiring patients to not have heart disease, while the criteria for trials using Convalescent Plasma Transfer and Anakinra are more lenient. Most drugs are given suitable scores for only a few severe patients. This demonstrates that AI4CoV recognizes that each patient will require different ranges of treatment options based on their underlying conditions.

As a validation, we inspected AI4CoV’s suitability predictions to 216 pairs of patient-trial scores consisting of the 9 trials of the drugs mentioned above and the 24 patients and found all of the predictions to be correct. This instance of AI4CoV was trained only with patient-trial pairs of the 28 mild and moderate patients from Taizhou and never saw any severe patients but still generalized well for severe patients. Some of the trials were used to pair with the mild and moderate patients in training. The eligibility criteria of these 9 clinical trials are relatively well-written compared to those of randomly selected COVID-19 clinical trials registered in ClinicalTrials.gov because they were sponsored by highly regarded institutions. The well-written criteria are less ambiguous for AI4CoV to reason and assign suitability scores than randomly selected COVID-19 clinical trials used in our performance evaluation.

## Limitations

AI4CoV’s suitability scores only assess whether a patient is eligible for a clinical trial based on the patient’s underlying conditions. It is important to note that the scores do not measure how likely if the treatment from a trial would be effective at curing the patient. Evaluation by the healthcare team is essential. Regulations about using drugs under clinical trials must be strictly followed.

## Conclusions

AI4CoV is a successful demonstration that it is possible to have AI predict if a patient is suitable or unsuitable for a clinical trial and therefore able to consider the drug in the trial as a treatment option. Our results indicate that searching for suitable treatment options from a large number of trials registered in ClinicalTrials.gov can be performed accurately. Patients would benefit from the treatment options identified as suitable or unsuitable for them during a pandemic when no treatment has been proven to be safe and effective for patients with various pre-conditions.

AI4CoV can be trained to read the clinical trial information of any diseases, which can then be used to match clinical trials for patients or vice versa. Potential use cases of AI4CoV include allowing clinical trial sponsors to rapidly search for eligible subjects if a database of candidate patients is available. This is particularly useful for the clinical trials of rare diseases and late-stage cancers that target a very specific population that has, for example, certain mutations present.^32^ AI4CoV can also help inform patients about clinical trials in which they may participate.

## Data Availability

The patient data for Lausanne University Hospital are available online (see citation 16). All of the clinical trials used to test our system are online in clinicaltrials.gov. The Taizhou patient data is not available online.

https://clinicaltrials.gov

http://doi.org/10.5281/zenodo.3763421

## Competing Interests

The authors whose names are listed immediately below certify that they have NO affiliations with or involvement in any organization or entity with any financial interest (such as honoraria; educational grants; participation in speakers’ bureaus; membership, employment, consultancies, stock ownership, or other equity interest; and expert testimony or patent-licensing arrangements), or non-financial interest (such as personal or professional relationships, affiliations, knowledge or beliefs) in the subject matter or materials discussed in this manuscript.

Author names: DongQing Lv, Jane Y-J Hsu, Pai Jung Huang.

The authors whose names are listed immediately below report the following details of affiliation or involvement in an organization or entity with a financial or non-financial interest in the subject matter or materials discussed in this manuscript.

Author names: Andrew Hsu, Amber Yeh, Shao-Lang Chen, Jerry Yeh.

These authors are employees of the AI4WARD Inc. which may potentially benefit from the research results of this study.

All the authors agree that the above information is true and correct.

## Author Contribution

All persons who meet authorship criteria are listed as authors, and all authors certify that they have participated sufficiently in the work to take public responsibility for the content, including participation in the concept, design, analysis, writing, or revision of the manuscript. Furthermore, each author certifies that this material or similar material has not been and will not be submitted to or published in any other publication. Authorship contributions

Conception and design of study: YJ Hsu, PJ Huang;

Acquisition of data: D Lv, PJ Huang.

Analysis and/or interpretation of data: A Hsu, A Yeh, D Lv, PJ Huang.

Drafting the manuscript: A Hsu, A Yeh;

Revising the manuscript critically for important intellectual content: SL Chen, J Yeh, YJ Hsu, PJ Huang.

Approval of the version of the manuscript to be published (the names of all authors must be listed): A Hsu, A Yeh, SL Chen, J Yeh, D Lv, YJ Hsu, PJ Huang.

We have not received substantial contributions from non-authors.

## Data Availability

The patient data for Lausanne University Hospital are available online (see citation 16). All of the clinical trials used to test our system are online in ClinicalTrials.gov. The Taizhou patient data is not available online.

